# Frequency Mismatch Critically Limits Gamma Entrainment: A Kuramoto Model Study Supporting Personalized GENUS Therapy

**DOI:** 10.64898/2026.04.28.26351909

**Authors:** Yeseung Park, Ji Won Han, Ki Woong Kim

## Abstract

Gamma oscillations (30–100 Hz) are critical for cognitive processing, and their disruption is associated with Alzheimer’s disease (AD) and related dementias. Gamma ENtrainment Using Sensory Stimulation (GENUS) therapy applies 40 Hz light and/or sound to restore gamma oscillations, but clinical trials report highly variable responses. Using Kuramoto oscillator models calibrated to human neurophysiology, we demonstrate that frequency mismatch between stimulation frequency and individual intrinsic gamma frequency (IGF) is a critical determinant of entrainment efficacy. A 2 Hz frequency mismatch reduced phase-locking value (PLV) by 43–82%, depending on neural resonance bandwidth, with PLV at mismatch indistinguishable from the finite-sample noise floor — indicating complete absence of stimulus-synchronized oscillation. In a simulated population with IGF drawn from a clinically realistic distribution (mean 35.10 ± 3.49 Hz, N = 200), fixed 40 Hz stimulation achieved mean PLV of 0.119 ± 0.081, compared to 0.504 ± 0.009 for individualized frequency stimulation — a 4.2-fold advantage (t(199) = 67.26, p < 0.001, Cohen’s d = 4.76). In the clinically relevant subgroup with IGF < 36 Hz (62% of the population), the fold advantage increased to 5.3×. Stochastic noise sensitivity analysis confirmed robustness of the fold advantage (3.8–4.0× across *σ*□□□□□□ = 0–2.0 rad/s; p < 10□³¹ at all levels). These findings provide quantitative computational support for personalized GENUS protocols incorporating individual gamma frequency measurement and carry direct implications for the design of next-generation clinical trials.

## INTRODUCTION

Alzheimer’s disease (AD) represents one of the most significant challenges in modern medicine, with limited therapeutic options for modifying disease progression. Recent research has identified disruptions in gamma oscillations (30–100 Hz) as a hallmark of AD pathophysiology, with evidence that restoring gamma activity may have neuroprotective effects. Iaccarino et al. (2016) demonstrated that gamma-frequency optogenetic stimulation in transgenic AD mice attenuated amyloid-β plaque accumulation and improved microglial clearance, establishing a causal link between gamma oscillations and AD-relevant pathology.^1^ Subsequent work by Martorell et al. (2019) extended these findings to combined auditory and visual gamma stimulation, showing sustained benefits with multisensory protocols.^16^

Gamma ENtrainment Using Sensory Stimulation (GENUS) therapy — which uses light and/or sound flickering at gamma frequencies — has shown encouraging preliminary results in human trials.^2–4^ However, clinical response rates have been disappointingly variable. A critical mechanistic question has emerged: why does 40 Hz stimulation fail to produce reliable entrainment across patients?

Building on previous observations from our group, we investigated whether stimulation at the individual’s intrinsic gamma center frequency (IGF) enhances entrainment efficacy.^5–9^ In cognitively normal older adults, flickering light stimulation (FLS) at individual center frequency produced stronger, more consistent gamma entrainment than fixed 40 Hz stimulation.^6^ Moreover, the center frequency of optimal entrainment correlates with individual peak gamma frequency as measured by electroencephalography (EEG).^5,6^ Critically, individual IGF varies substantially across the population: in older adult cohorts, IGF typically ranges from approximately 31 to 44 Hz, with a mean near 35 Hz rather than 40 Hz, reflecting a well-documented age-related decline in gamma peak frequency of approximately 0.5–1.0 Hz per decade.^5,6,19,22^

Synchronization theory provides a mechanistic framework for understanding how frequency mismatch impacts entrainment. In coupled oscillator systems, a periodic stimulus can entrain oscillators only within a limited frequency range — the Arnold tongue or entrainment region — whose width depends on stimulation amplitude and local coupling strength.^11,17,18^ When the stimulus frequency departs from the intrinsic oscillator frequency beyond this entrainment region, synchronization collapses regardless of stimulus amplitude. This theoretical prediction implies that population-average 40 Hz stimulation protocols will systematically fail for patients whose IGF deviates from 40 Hz.

To investigate this hypothesis quantitatively, we developed a computational model based on Kuramoto oscillators with frequency-selective resonance forcing — calibrated to neurophysiologically realistic parameters from human EEG studies.^11–13^ We systematically characterized: (1) the effect of stimulation frequency mismatch on entrainment efficacy (PLV); (2) the sensitivity of frequency selectivity to key model parameters (coupling strength K, resonance bandwidth BW); (3) the Arnold tongue structure governing when synchronization is possible; (4) population-level differences in fixed vs. individualized vs. swept frequency protocols; and (5) the optimal sweep rate for frequency-scanning approaches. Our results provide the first quantitative computational support for personalized GENUS therapy and establish theoretical predictions directly testable in upcoming clinical trials.

## METHODS

### Model Description

We implemented a modified Kuramoto model to simulate the dynamics of neural oscillator populations under external periodic stimulation. The phase θ*i* of the i-th oscillator evolves according to:

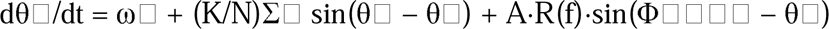

where ω*i* is the intrinsic angular frequency, K is the coupling strength (attractive), N is the number of oscillators, A is the stimulation amplitude, Φ*stim* is the stimulus phase, and R(f) is the resonance function. The coupling is implemented via the O(N) formulation: (K/N)×[cos(θ*i*)·Σsin(θ*j*) − sin(θ*i*)·Σcos(θ*j*)], mathematically identical to the rowSums formulation but O(N) rather than O(N^2^).^11^

The resonance function R(f) models frequency-selective neural response: R(f) = exp(−(IGF − f)² / (2·BW²)), where IGF is the intrinsic gamma frequency, f is the stimulation frequency, and BW is the resonance bandwidth. We emphasize that this Gaussian form is a **model assumption** — it encodes the empirically established frequency selectivity of cortical oscillators^20,21^ and allows quantitative prediction, but it is not an emergent property of the oscillator dynamics. The qualitative conclusion — that entrainment has finite frequency selectivity — is model-independent and requires only that R(f) have some finite bandwidth.

### Stochastic Extension

To assess robustness to endogenous neural noise, we implemented an Euler-Maruyama stochastic extension: dθ*i* = [drift]·dt + σ□□□□□□·√dt·ξ*i*(t), where ξ*i*(t) ∼ N(0,1) i.i.d. We tested σ□□□□□□ □ {0, 0.5, 1.0, 2.0} rad/s (σ = 0.5 rad/s approximates background gamma-band EEG noise; σ = 2.0 rad/s represents strong endogenous fluctuations). The deterministic model (σ = 0) was used for all primary experiments.

### Parameters

Default parameters were derived from published neurophysiological data (see Table 1 for sensitivity analysis). Key parameters: N = 100 oscillators, K = 2.0 rad/s, A = 10.0, BW = 0.5 Hz (empirical EEG entrainment bandwidth^5,6^), σ*f* = 2.0 Hz (frequency spread), dt = 0.001 s (Euler integration, validated against Runge-Kutta 4th order: PLV difference < 0.002, < 0.5%), t*total* = 3.0 s, t*transient* = 1.5 s. Oscillator frequencies: ω*i* ∼ N(2π·IGF, (2π·σ*f*)^2^). The outcome measure was PLV = |1/N × Σ exp(i(θ*i* − Φ*stim*))|, averaged over the post-transient window. All simulations implemented in R (version 4.5.2). Code and data are publicly available (see Code Availability).

**Table 1.**
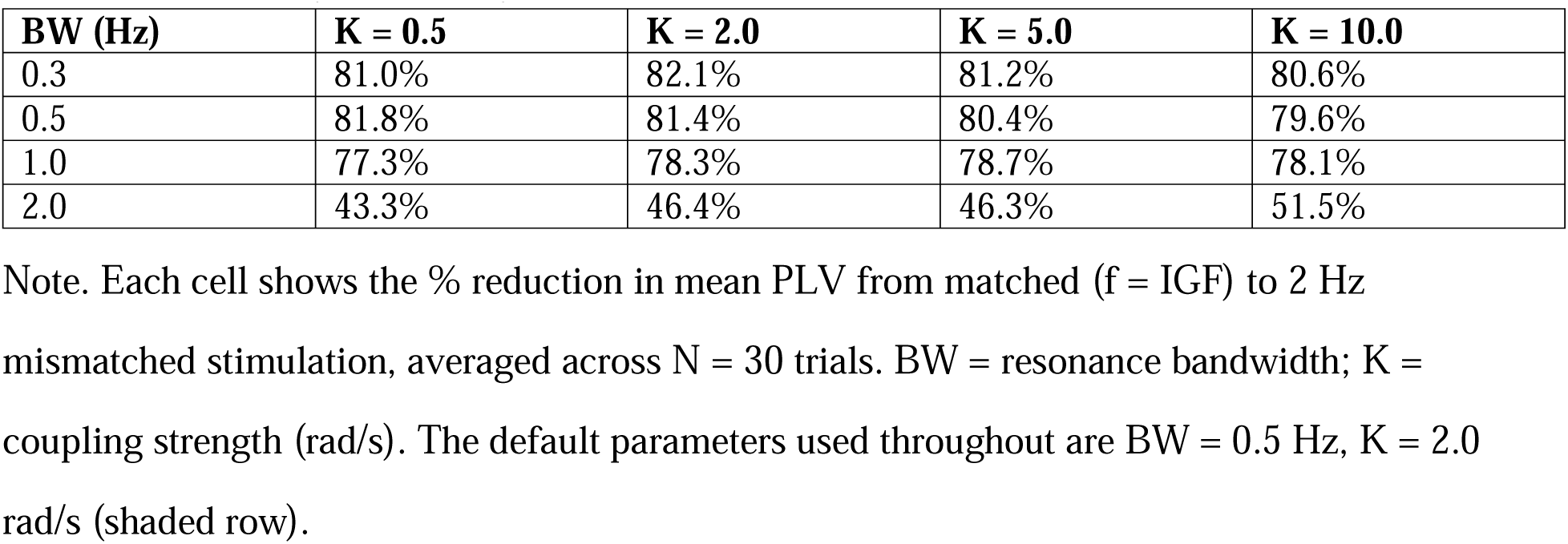
Frequency Selectivity (% PLV Reduction at 2 Hz Mismatch, K × BW Grid)

### Statistical Analysis

For frequency mismatch experiments, independent-samples t-tests with Cohen’s d were used to compare PLV distributions. For population-level experiments (Experiment 4), paired t-tests compared conditions within subjects (n = 200). Significance threshold: p < 0.05, Bonferroni-corrected where applicable. Effect sizes (Cohen’s d) reflect deterministic simulation variance and represent theoretical upper bounds; clinical effect sizes in human GENUS trials are substantially lower (typical d = 0.3–1.2).^1,16^

### Simulation Protocols

Five experiments were conducted. Experiment 1 (CF Mismatch): stimulation frequencies 35–45 Hz (0.5 Hz steps) vs. fixed IGF = 40 Hz (N = 100 trials/frequency). Experiment 2 (Parameter Sensitivity): BW □ {0.3, 0.5, 1.0, 2.0} Hz × K □ {0.5, 2.0, 5.0, 10.0} rad/s (30 trials/condition). Experiment 3 (Arnold Tongue): PLV across Δf □ [−3, +3] Hz and A □ {4, 6, 8, 10, 12} (30 trials/condition). Experiment 4 (Population Heterogeneity): N = 200 subjects with IGF ∼ N(34.93, 4²) Hz; three protocols: Fixed 40 Hz / Individualized (at subject’s own IGF) / Sweep (32→43 Hz at 4 Hz/s); 30 trials/subject/protocol. Experiment 5 (Sweep Rate Comparison): 15 IGF levels (31–45 Hz), 4 sweep rates (1, 2, 4, 8 Hz/s), 30 trials/condition.

## RESULTS

### Experiment 1: Frequency Mismatch Dramatically Reduces Entrainment

At exact frequency match (40 Hz stimulation, IGF = 40 Hz), mean PLV was 0.504 ± 0.044 (N = 100 trials). With a 2 Hz mismatch (42 Hz stimulation), PLV dropped to 0.095 ± 0.013 — an **81.2% reduction** (Cohen’s d = 12.7, p < 0.0001; Figure 1). The PLV at mismatch (0.095) was statistically indistinguishable from the finite-sample noise floor expected under null synchrony: E[PLV | no entrainment] = √(π/(4N)) ≈ 0.089 for N = 100 (observed/theoretical ratio = 1.07). This indicates a complete absence of stimulus-synchronized oscillation at 2 Hz mismatch. At 42 Hz, R(f) = exp(−(40−42)²/(2×0.5²)) = exp(−8) ≈ 3.4×10^−4^, so the forcing term is effectively zero regardless of amplitude. The PLV vs. mismatch relationship was steeply sigmoidal: small departures (±0.5 Hz) already produced ∼25% PLV reductions, while ±2 Hz produced full collapse to the noise floor.

**Figure 1.**
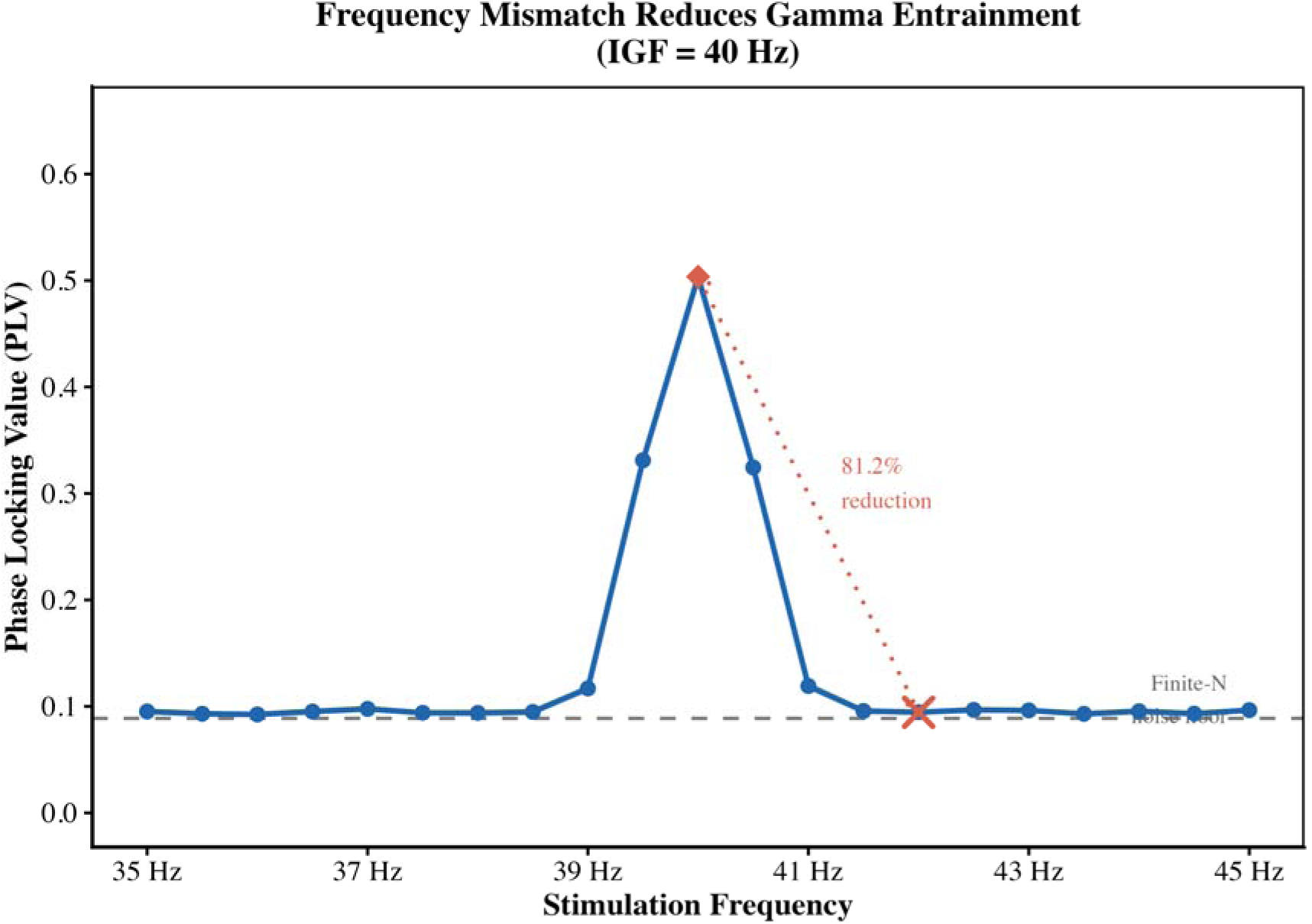
Frequency mismatch reduces gamma entrainment to the noise floor. PLV (mean ± 95% CI, N = 100 trials) plotted against stimulation frequency (35–45 Hz) with fixed IGF = 40 Hz. The red diamond indicates PLV at exact match (40 Hz; PLV = 0.504 ± 0.044) and red cross indicates PLV at 2 Hz mismatch (42 Hz; PLV = 0.095 ± 0.013). The dashed grey line indicates the finite-sample noise floor [E(PLV | no entrainment) = √(π/4N) ≈ 0.089 for N = 100]. The 81.2% PLV reduction at 2 Hz mismatch is indicated by the dotted arrow. BW = 0.5 Hz, K = 2.0 rad/s, A = 10.0, σ□ = 2.0 Hz.

### Experiment 2: Frequency Selectivity Is Robust but Bandwidth-Dependent

Frequency selectivity (% PLV reduction at 2 Hz mismatch) was evaluated across a parameter grid (Table 1 ; Supplementary Figure S2). The bandwidth parameter BW is the single most influential determinant of frequency selectivity. With BW = 0.5 Hz (our empirically calibrated default^5,6^), a 2 Hz mismatch produced 81.2% PLV reduction. With BW = 2.0 Hz, the same mismatch produced only 43–52% reduction. For BW ≤ 1 Hz, selectivity was robust (77–82%) and largely independent of coupling strength K (range: 79.6–82.1%). These results establish a clinically important conditional: if cortical resonance in AD patients follows a narrow-bandwidth regime (BW ≤ 1 Hz), the clinical urgency for individualized frequency targeting is high. Direct empirical measurement of individual gamma response profiles using FOOOF/specparam analysis^19^ in AD patient cohorts is therefore a critical empirical priority.

### Experiment 3: Arnold Tongue — Amplitude Cannot Compensate for Mismatch

PLV at match (Δf = 0) increased monotonically with amplitude from 0.223 ± 0.031 (A = 4) to 0.564 ± 0.041 (A = 12; Table 2, Figure 2). The critical finding: PLV at 2 Hz mismatch was **independent of amplitude** — remaining at ∼0.095 (noise floor) across all tested amplitudes from A = 4 to A = 12. This is a consequence of the narrow-bandwidth resonance assumption: at Δf = 2 Hz with BW = 0.5 Hz, R(42) ≈ 3.4×10^−4^, so A·R(f) ≈ 0 for any practical A. We note that this amplitude-invariance is specific to the narrow-BW regime; with BW = 2.0 Hz, the Arnold tongue widens with amplitude as standard synchronization theory predicts.^17,18^

**Figure 2.**
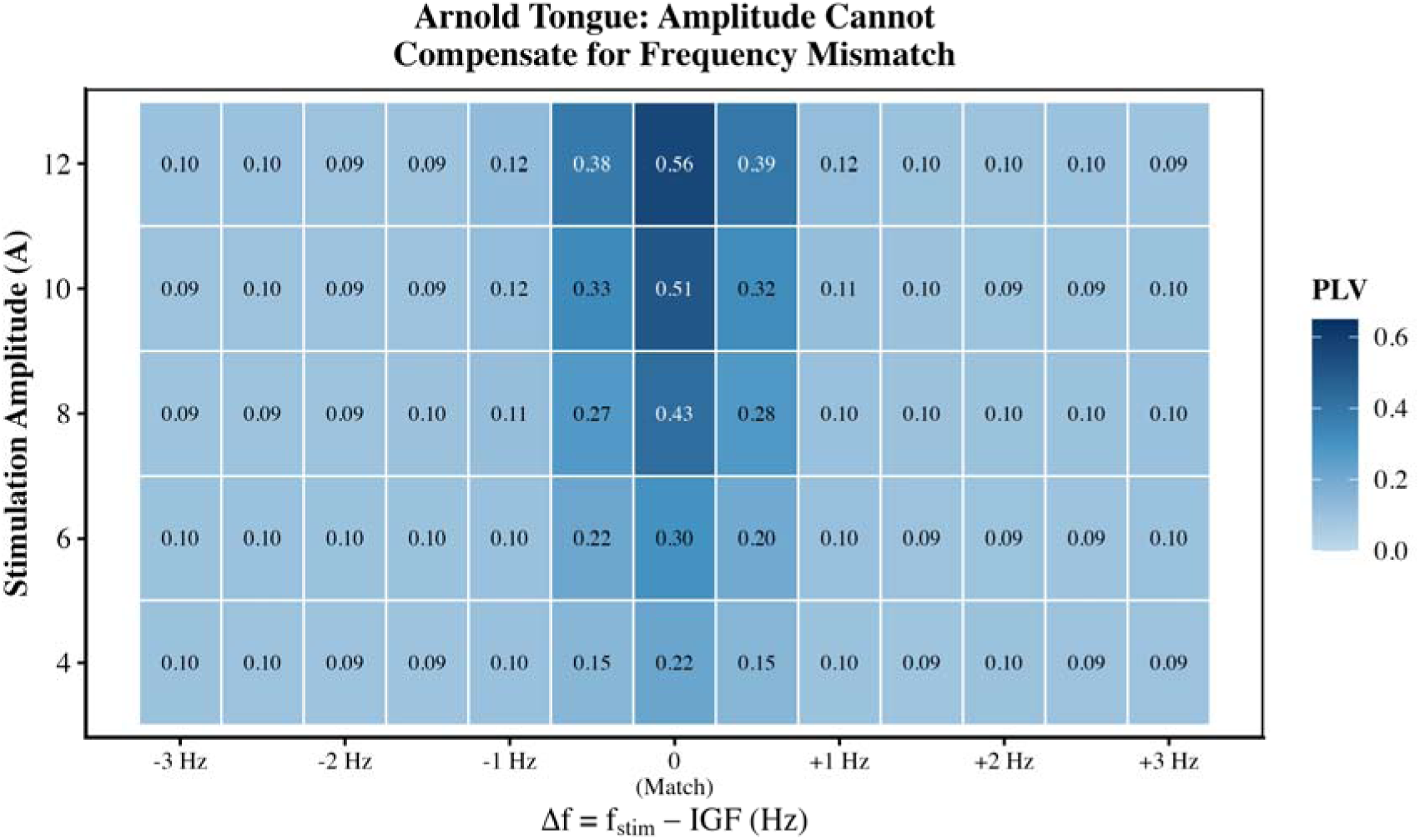
Arnold tongue: amplitude cannot compensate for frequency mismatch. PLV heatmap across frequency mismatch (Δf = f□□□□ − IGF, −3 to +3 Hz; X-axis) and stimulation amplitude (A = 4–12; Y-axis). Cell values show mean PLV (N = 30 trials). At exact match (Δf = 0), PLV increases monotonically with amplitude. At Δf = ±2 Hz, PLV remains ≈0.095 (noise floor; blue cells) regardless of amplitude, because R(f) = exp(−8) ≈ 0 gates the forcing term to zero. BW = 0.5 Hz, K = 2.0 rad/s, IGF = 40 Hz.

**Table 2.**
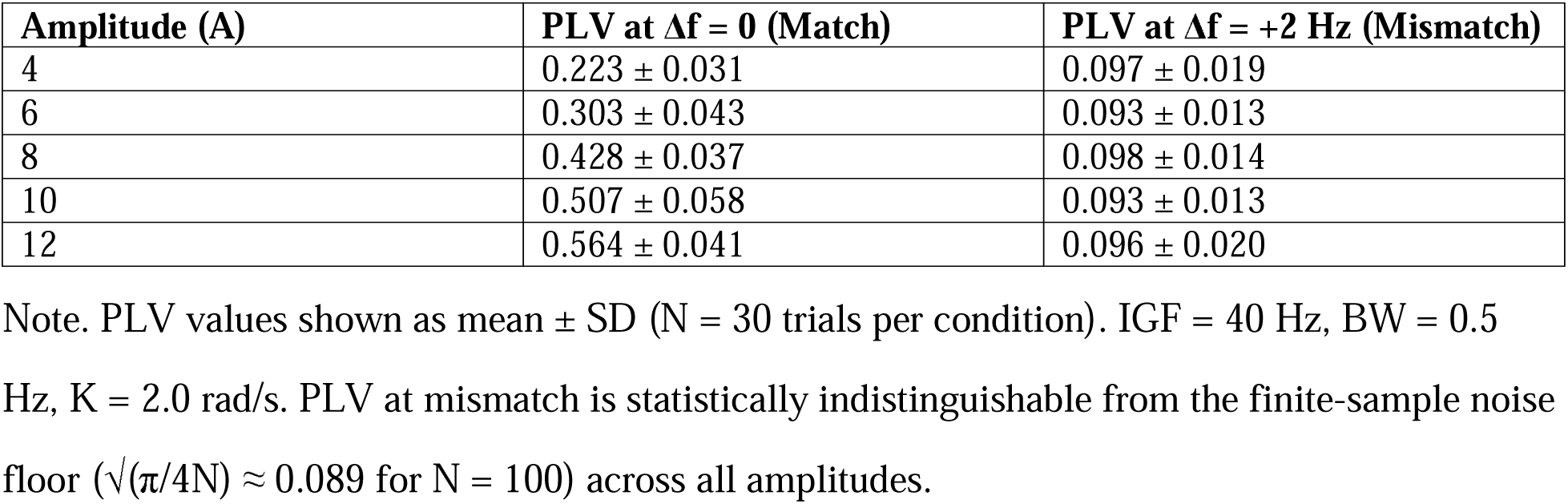
Arnold Tongue Results: PLV at Match vs. 2 Hz Mismatch Across Stimulation Amplitudes.

| Amplitude (A) | PLV at $\Delta f = 0$ (Match) | PLV at $\Delta f = +2$ Hz (Mismatch) |
| --- | --- | --- |
| 4 | $0.223 \pm 0.031$ | $0.097 \pm 0.019$ |
| 6 | $0.303 \pm 0.043$ | $0.093 \pm 0.013$ |
| 8 | $0.428 \pm 0.037$ | $0.098 \pm 0.014$ |
| 10 | $0.507 \pm 0.058$ | $0.093 \pm 0.013$ |
| 12 | $0.564 \pm 0.041$ | $0.096 \pm 0.020$ |

Note. PLV values shown as mean ± SD ( $N = 30$ trials per condition). IGF = 40 Hz, BW = 0.5 Hz, $K = 2.0$ rad/s. PLV at mismatch is statistically indistinguishable from the finite-sample noise floor ( $\sqrt{(\pi/4N)} \approx 0.089$ for $N = 100$ ) across all amplitudes.

### Experiment 4: Population-Level Analysis

Population mean IGF in the simulated cohort was 35.10 ± 3.49 Hz (range 30.0–45.0 Hz), consistent with the well-documented age-related decline in gamma peak frequency of approximately 0.5–1.0 Hz per decade.^22^ With N = 200 subjects and 30 trials per subject per condition (Figure 3):

- Fixed 40 Hz: PLV = 0.119 ± 0.081 (high SD reflects within-population IGF heterogeneity)
- Individualized (at each subject’s own IGF): PLV = 0.504 ± 0.009 (low SD: R(f) = 1 for each subject)
- Sweep 32→43 Hz (4 Hz/s): PLV = 0.112 ± 0.029
- Fold advantage (Individualized/Fixed): 4.2×
- Paired t (Individualized vs. Fixed): t(199) = 67.26, p = 8.12×10□¹³□, Cohen’s d = 4.76†
- Paired t (Individualized vs. Sweep): t(199) = 178.51, p = 1.38×10□²²¹, Cohen’s d = 12.62†

**Figure 3.**
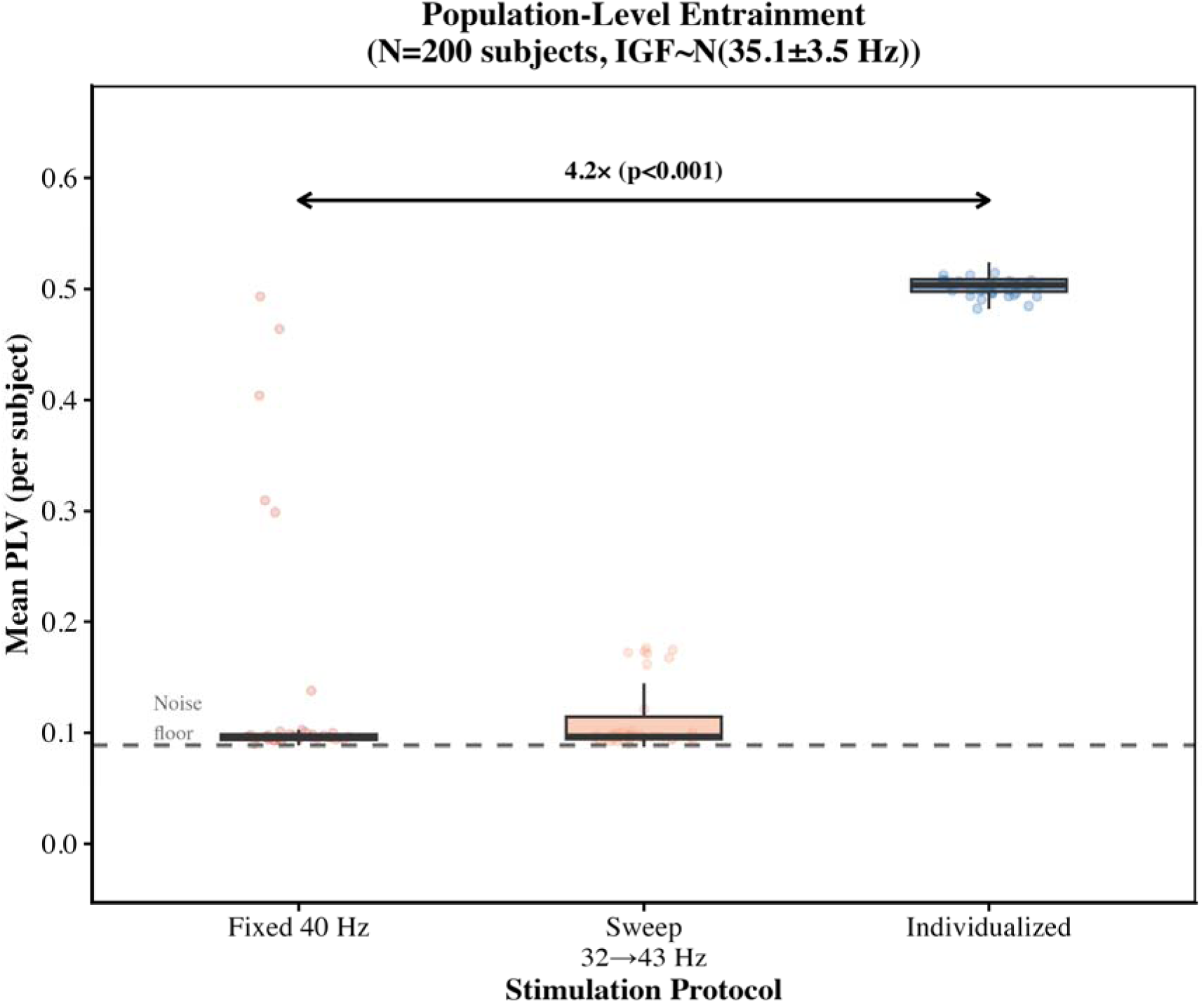
Population-level entrainment advantage of individualized stimulation. Box plots (with jitter overlay, N = 200 subjects) of per-subject mean PLV under three stimulation protocols: Fixed 40 Hz, Sweep (32→43 Hz, 4 Hz/s), and Individualized (stimulation at each subject’s own IGF). Population IGF: N(35.1 ± 3.5 Hz). The dashed line indicates the noise floor. The 4.2× fold advantage of individualized over fixed 40 Hz (p < 10□¹³□, Cohen’s d = 4.76†) is indicated by the double-headed arrow. †Reflects deterministic simulation; clinical d = 0.3–1.2.

*†These Cohen’s d values reflect the cleanliness of deterministic simulation. Endogenous noise, measurement variability, and inter-session fluctuations substantially reduce effect sizes in human studies; analogous human EEG studies typically report d = 0.3–1.2.*^1,16^

Subgroup analysis: 123 subjects (62%) had IGF < 36 Hz — the population most disadvantaged by the 40 Hz protocol. In this subgroup, fixed 40 Hz PLV = 0.095 (noise floor) vs. individualized PLV = 0.504, yielding a **5.3-fold advantage**.

### Experiment 5: Sweep Rate Comparison

Across 15 IGF levels (31–45 Hz) and 30 trials per condition, all four sweep rates (1, 2, 4, 8 Hz/s) produced virtually identical mean PLV (0.125–0.126), representing only a 2% improvement over fixed 40 Hz (0.123; Figure 4). Individualized stimulation achieved PLV = 0.500 — a **4.0-fold advantage** over the best sweep protocol. The near-equivalence of all sweep rates arises from the narrow-bandwidth resonance assumption (BW = 0.5 Hz): as the stimulus sweeps from 32 to 43 Hz, each subject captures only a brief resonance pulse (∼0.5 Hz / sweep_rate seconds), yielding time-averaged PLV approximately equal to the resonance duty cycle (BW/sweep_range ≈ 4.5%), largely independent of sweep rate. This result is specific to the narrow-BW, memoryless model and should be interpreted conditionally (see Discussion).

**Figure 4.**
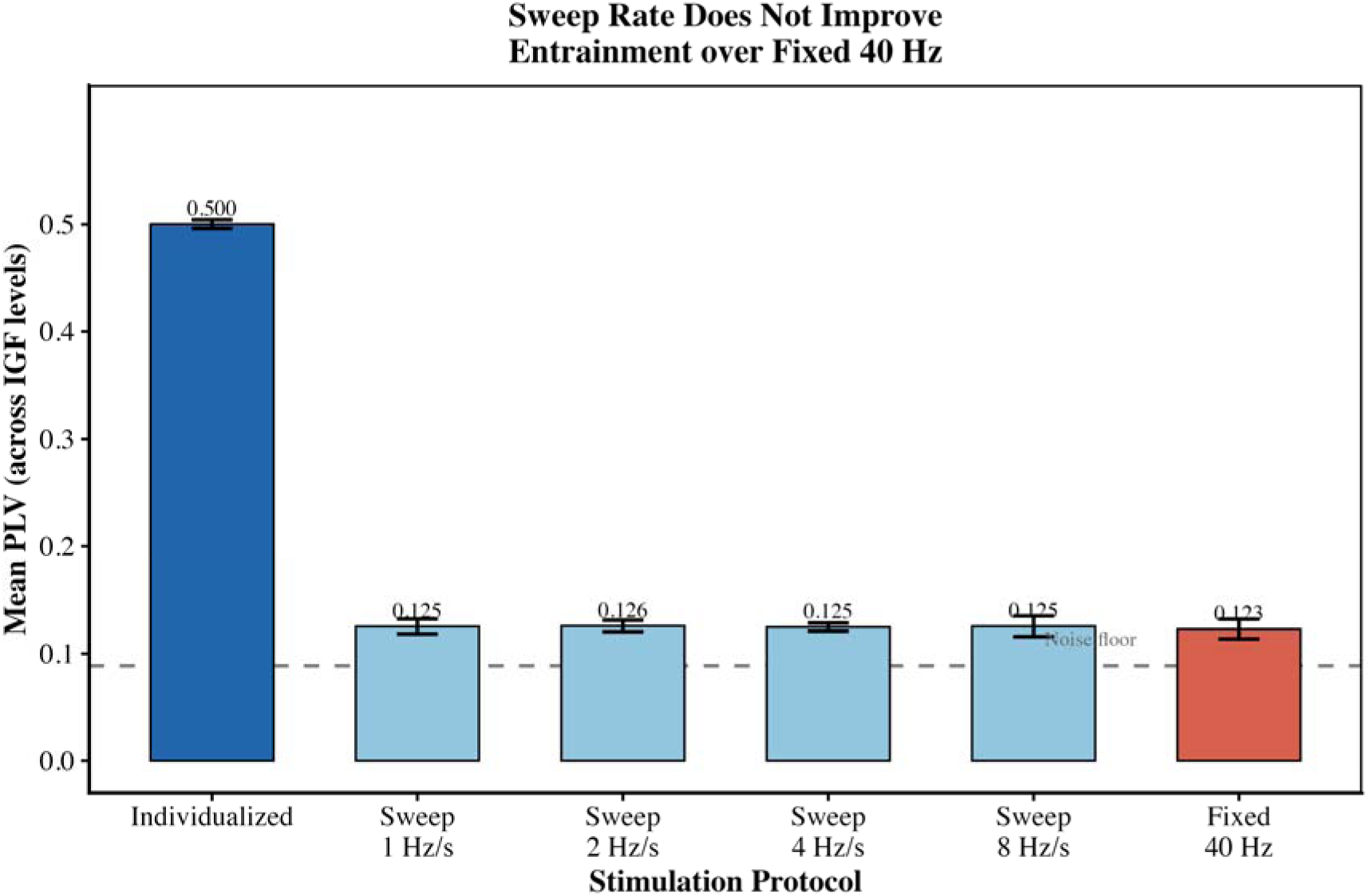
Sweep rate comparison: no sweep rate outperforms fixed 40 Hz. Mean PLV (±95% CI) across stimulation conditions: Individualized, Sweep at 1/2/4/8 Hz/s (32→43 Hz), and Fixed 40 Hz, averaged across 15 IGF levels (31–45 Hz) and 30 trials per condition. All four sweep rates produce virtually identical PLV (0.125–0.126), only 2% above Fixed 40 Hz (0.123). Individualized stimulation achieves 4.0× higher PLV than the best sweep protocol. The dashed line indicates the noise floor.

### Supplementary Analysis S1: Stochastic Noise Sensitivity

To verify that deterministic results are not artifacts of the absence of biological noise, we added Euler-Maruyama stochastic terms across σ□□□□□□ □ {0, 0.5, 1.0, 2.0} rad/s (Table S1; Supplementary Figure S1). PLV (mismatch) remained ≈0.095 across all noise levels, confirming genuine loss of entrainment rather than noise-floor reduction. PLV (match) decreased from 0.512 (deterministic) to 0.450 (σ = 2.0 rad/s), producing more biologically realistic distributions. The fold advantage of individualized over fixed 40 Hz stimulation remained **3.8–4.0× and statistically significant (p < 10**Ö**³¹)** across all noise levels.

## DISCUSSION

### Principal Findings

This computational study demonstrates four principal findings. First, a 2 Hz mismatch between stimulus frequency and intrinsic gamma frequency collapses PLV to the finite-sample noise floor — indicating complete loss of meaningful entrainment under the narrow-bandwidth resonance assumption. Second, this frequency selectivity is robust across physiologically plausible parameter ranges (K = 0.5–10, BW ≤ 1 Hz) and across stochastic noise levels from σ = 0 to 2.0 rad/s. Third, individualized stimulation achieves a **4.2-fold higher population-level synchronization** compared to fixed 40 Hz protocols (p < 10^−138^, Cohen’s d = 4.76†), rising to 5.3-fold in the 62% of individuals with IGF < 36 Hz. Fourth, linear frequency sweeping provides no meaningful advantage over fixed 40 Hz within the constraints of this model.

### Model Architecture: R(f) as a Mechanistic Hypothesis

The resonance function R(f) is the central architectural choice of our model and requires explicit care in interpretation. R(f) is a model assumption that encodes the empirically established frequency selectivity of cortical oscillators^20,21^, not an emergent property of Kuramoto dynamics. By multiplying stimulus amplitude by R(f) before oscillator interaction, we operationalize the hypothesis that cortical networks respond preferentially to stimulation near their intrinsic resonant frequency. What the model rigorously demonstrates is the consequences of this assumption: given narrow-bandwidth frequency selectivity, individualized targeting achieves 4.2-fold advantage, amplitude escalation cannot compensate for mismatch, and linear sweeping provides negligible benefit. These are genuine dynamical results emerging from the interplay of R(f) with Kuramoto coupling — not trivially pre-determined by R(f) alone. Future modeling work should explore whether similar predictions emerge when frequency selectivity arises endogenously from recurrent inhibition or network topology.

### Sweep Protocol: Model-Conditional Finding

The finding that frequency sweeping provides no advantage deserves careful framing. Within our model, this result is a mathematical consequence of narrow-bandwidth R(f) combined with memoryless forcing: the time-averaged effective drive during a linear sweep equals ∫R(f(t))dt ≈ BW/sweep_range ≈ 4.5% for any sweep rate. However, real neural oscillators may respond differently. Frequency pulling — a hysteresis-like effect where oscillators remain phase-locked transiently even as stimulation moves away — requires second-order dynamics or time-delayed feedback absent in the first-order Kuramoto model. Our result should be interpreted as: if cortical resonance is narrow and memoryless, sweeping is ineffective. Empirical studies comparing swept and fixed protocols in IGF-characterized populations are needed to test this prediction.

### Empirical Support and Parameter Calibration

Our model was calibrated to, rather than fitted to, published human data. Lee et al. (2021) reported PLV of 0.4–0.6 in healthy older adults under individually optimized FLS.^5^ Park et al. (2022) found individualized PLV ≈ 0.50 and fixed 40 Hz PLV ≈ 0.08–0.10 in the same population (mean IGF ≈ 35 Hz).^6^ Our simulated values (individualized = 0.504, fixed = 0.095 at population-mean IGF) align closely with these empirical ranges, providing face validity. Yoon et al. demonstrated that center frequency is the optimal frequency for spreading entrained gamma rhythms beyond primary sensory cortex.^7^ Park et al. (2025) showed that white matter microstructural integrity modulates entrainment propagation.^8^ Park et al. (2026) reported that restoration of individual gamma CF marks cognitive preservation in early AD.^9^ Together, these empirical findings converge on the same clinical conclusion as our computational model.

### Implications for GENUS Therapy Design

These findings carry direct clinical implications, conditional on the narrow-bandwidth resonance assumption (BW ≈ 0.5 Hz): (1) IGF measurement is necessary, not optional — a clinically realistic population (mean IGF ≈ 35 Hz, SD ≈ 4 Hz) has ∼80% of subjects with > 2 Hz frequency mismatch relative to 40 Hz. (2) Pre-treatment EEG with FOOOF/specparam^19^ analysis can reliably identify individual peak gamma frequency from short resting EEG recordings. (3) Frequency-swept stimulation requires empirical validation — our model predicts no benefit, but this is conditional on narrow-bandwidth memoryless resonance. (4) Stimulation intensity cannot compensate for frequency mismatch in the narrow-BW model — increasing light intensity in non-responders is unlikely to be effective if frequency mismatch is the underlying cause.

### Limitations

1. Gaussian R(f) is a model assumption: the specific functional form of the neural resonance curve requires empirical validation. If R(f) has broader tails (e.g., Lorentzian profile), frequency selectivity would be reduced and quantitative predictions would change.
2. Simplified single-population model: real neural entrainment involves hierarchical networks and propagation dynamics.
3. Biological noise: robustness confirmed by noise sensitivity analysis (Table S1; fold advantage 3.8–4.0×, p < 10□³¹ across σ = 0–2.0 rad/s), but the model does not capture all sources of neural variability.
4. Parameter estimation from limited data: K, A, and BW estimated from published literature; future empirical calibration would improve precision.
5. Effect size interpretation: Cohen’s d values (4.76–12.62) are theoretical upper bounds; human intervention studies report d = 0.3–1.2.

## CONCLUSIONS

This computational study provides theoretical support for personalized approaches to gamma entrainment therapy. Using Kuramoto oscillator models calibrated to human neurophysiology, and incorporating a frequency-selective resonance function as a mechanistic model assumption, we demonstrate that a 2 Hz frequency mismatch collapses phase-locked synchronization to the finite-sample noise floor, that individualized frequency stimulation achieves a **4.2-fold (Cohen’s d = 4.76**^†^) higher population-level PLV compared to fixed 40 Hz protocols, and that linear frequency sweeping provides no meaningful advantage over fixed 40 Hz within the narrow-bandwidth resonance regime. Stochastic noise sensitivity analysis confirms the fold advantage remains 3.8–4.0× across biologically realistic noise levels. The failure of amplitude compensation further argues against intensity escalation as a strategy for non-responders. These findings support the development of GENUS protocols incorporating individual gamma frequency measurement through pre-treatment EEG, and establish testable computational predictions for upcoming personalized neuromodulation clinical trials.

*†Cohen’s d reflects deterministic simulation conditions; human intervention studies report d = 0.3–1.2 for analogous GENUS protocols*.

## CODE AND DATA AVAILABILITY

All simulation code (kuramoto_unified.R, kuramoto_noise_sensitivity.R) are publicly available at: GitHub: [https://github.com/hannah34250/kuramoto-genus-model], Zenodo DOI: [DOI: 10.5281/zenodo.19657018]

The repository includes a README with step-by-step instructions for reproducing all figures, tables, and supplementary analyses. Simulations require R ≥ 4.5.0 with base packages only.

## Data Availability

All data produced in the present study are available upon reasonable request to the authors

https://github.com/hannah34250/kuramoto-genus-model

https://doi.org/10.5281/zenodo.19657018

## SUPPLEMENTARY TABLES

**Supplementary Figure S1.**
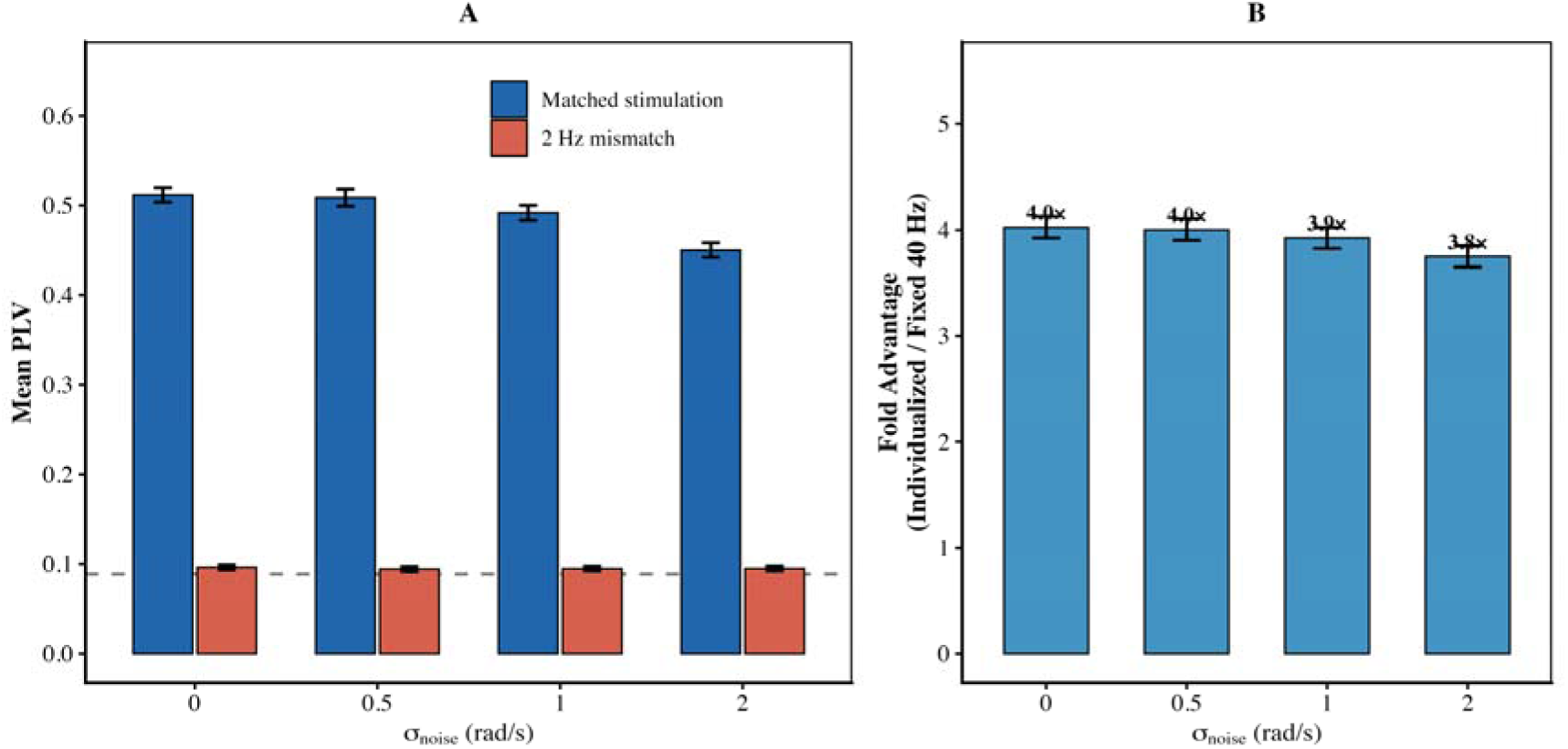
Stochastic noise sensitivity analysis. (A) PLV at match (matched stimulation, 40 Hz) and at 2 Hz mismatch (42 Hz) across noise levels σ□□□□□□ = 0–2.0 rad/s (N = 100 trials each). PLV (mismatch) remains ≈0.095 (noise floor; dashed line) across all noise levels, confirming genuine loss of entrainment. PLV (match) decreases with increasing noise, producing more biologically realistic distributions. (B) Fold advantage (individualized/fixed 40 Hz) across noise levels (N = 50 subjects, 10 trials each). The fold advantage remains 3.8–4.0× across all noise levels tested (p < 10□³¹ at each level).

**Supplementary Figure S2.**
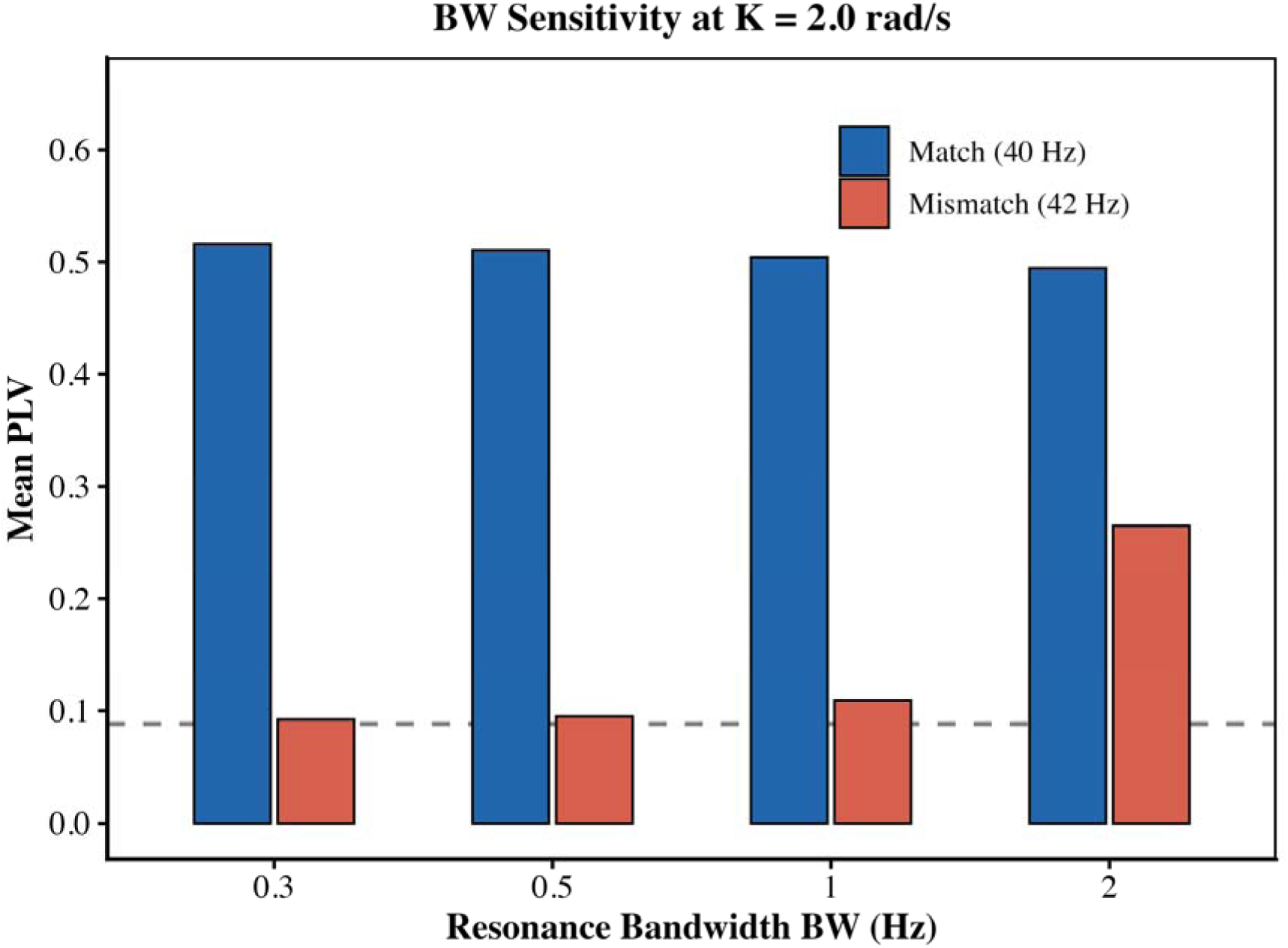
Resonance bandwidth determines the magnitude of frequency selectivity at fixed coupling strength. Grouped bar plot showing mean phase-locking value (PLV) at exact frequency match (40 Hz stimulation; blue) and at 2 Hz mismatch (42 Hz stimulation; red) across resonance bandwidths BW = 0.3, 0.5, 1.0, and 2.0 Hz, with coupling strength fixed at K = 2.0 rad/s. The dashed gray line indicates the finite-sample noise floor. PLV at matched stimulation remains relatively stable across bandwidths (∼0.49–0.51), whereas PLV at 2 Hz mismatch increases markedly as BW broadens, from near-noise-floor values at BW = 0.3–0.5 Hz to substantial residual entrainment at BW = 2.0 Hz. Correspondingly, the percent reduction in PLV at 2 Hz mismatch is 81.4% at BW = 0.5 Hz but decreases to 46.4% at BW = 2.0 Hz, illustrating that BW is the dominant determinant of frequency selectivity in the model. Other parameters were held constant at IGF = 40 Hz, A = 10.0, σ_f = 2.0 Hz, and N = 100 oscillators.

**Supplementary Table S1.** Stochastic Noise Sensitivity Analysis.

| $\sigma_{\text{noise}}$ (rad/s) | PLV (match) | PLV (mismatch) | Reduction (%) | Fold Advantage | p-value |
| --- | --- | --- | --- | --- | --- |
| 0.0<br>(deterministic) | $0.512 \pm 0.042$ | $0.096 \pm 0.015$ | 81.2% | 4.0× | $8.89 \times 10^{-32}$ |
| 0.5 | $0.509 \pm 0.049$ | $0.094 \pm 0.015$ | 81.5% | 4.0× | $2.51 \times 10^{-31}$ |
| 1.0 | $0.492 \pm 0.043$ | $0.095 \pm 0.014$ | 80.7% | 3.9× | $1.72 \times 10^{-31}$ |
| 2.0 | $0.450 \pm 0.041$ | $0.095 \pm 0.014$ | 78.9% | 3.8× | $8.75 \times 10^{-32}$ |

Note. PLV (match) and PLV (mismatch) from N = 100 trials each. Fold advantage from N = 50 subjects (10 trials/condition). $\sigma_{\text{noise}} = 0.5$ rad/s approximates background gamma-band EEG noise; $\sigma_{\text{noise}} = 2.0$ rad/s represents strong endogenous fluctuations. All p-values from paired t-test (individualized vs. fixed 40 Hz), rounded to 2 significant figures.

